# Breakthrough SARS-CoV-2 infections in immune mediated disease patients undergoing B cell depleting therapy

**DOI:** 10.1101/2022.02.21.22271289

**Authors:** Cassandra M Calabrese, Elizabeth Kirchner, Elaine M Husni, Brandon P Moss, Anthony J Fernandez, Yuxuan Jin, Leonard H Calabrese

**Affiliations:** Department of Rheumatic and Immunologic Diseases, Cleveland Clinic; Mellen Center for Multiple Sclerosis Treatment and Research, Cleveland Clinic; Department of Dermatology, Cleveland Clinic; Quantitative Health Sciences, Cleveland Clinic

**Author notes:** Corresponding author: Leonard H. Calabrese, D.O., Professor of Medicine, Cleveland Clinic Lerner, College of Medicine of Case Western Reserve University, RJ Fasenmyer Chair of Clinical Immunology, Department of Rheumatic and Immunologic Diseases, 9500 Euclid Avenue, Desk A50, Cleveland, Ohio 44195.

## Abstract

**Objectives:** Immunocompromised patients with immune mediated inflammatory diseases (IMIDs), undergoing therapy with B cell depleting agents are among the most vulnerable to experience severe COVID-19 disease as well as respond sub-optimally to SARS CoV-2 vaccines yet little is known about the frequency or severity of breakthrough infection in this population. We have analyzed a large cohort of vaccinated IMIDs patients undergoing B cell depleting therapy for the presence of breakthrough infection and assessed their outcomes.

**Methods:** Utilizing specific ICD codes the pharmacy records and COVID-19 registry at the Cleveland clinic were used to identify all patients with IMIDS treated with B cell depleting monoclonal antibodies who were vaccinated against SARs CoV-2 and experienced breakthrough infections. Each EMR record was hand-reviewed to extract clinical data including vaccine history, demographics, comorbidities, other therapies, details of B cell depleting therapy, and outcomes. Univariate and multivariable logistic/proportional-odds regression models were used to examine the risk factors for severe outcomes.

**Results:** Of 1696 IMIDS patients on B cell depleting therapies 74 developed breakthrough COVID-19. Outcomes were severe with 24 (35%) hospitalized, 11 (15%) patients requiring critical care and 6 (8 %) deaths. Monoclonal antibodies were used on an outpatient basis to treat 21 with only a single patient requiring hospitalization without oxygen support and no deaths.

**Conclusions:** In IMIDS patients on B cell depleting therapies breakthrough infections are frequent and associated with severe outcomes. Outpatient use of monoclonal antibody therapy was associated with enhanced clinical outcomes.

## Introduction

The deployment of vaccines to both prevent infection with SARS-CoV-2 as well as limit the severity of COVID-19 has proven efficacious for the general population. However, data on patients with underlying immunocompromising conditions, including those with immune mediated inflammatory diseases (IMIDs), suggest they have both an increased likelihood for developing breakthrough infections as well as for experiencing more severe outcomes despite full vaccination status ^1,2^.

While there is great heterogeneity in the capacity of specific immunosuppressive agents to limit the integrated immune response to both natural infection as well as vaccines, of particular concern is the class of B cell depleting therapies (BCDTs) widely used to treat an array of IMIDs. Before the introduction of vaccines, treatment of both rheumatic and neurologic IMIDs with such BCDTs was associated with more severe COVID-19 ^3–9^. Furthermore, numerous studies have documented the capacity for BCDTs such as rituximab and ocrelizumab to profoundly impair humoral response to numerous vaccines including SARS-CoV-2 ^10–13^. More recently, however, several studies ^14,15^ have documented a dichotomy in vaccine responses in IMIDS patients receiving BCDTs, demonstrating suppression of the humoral response while documenting a preserved and robust cell mediated immune response - raising hope that such a pattern of immunity will afford adequate protection. Until now there has been no systematic investigation of IMIDs patients on BCDTs examining the frequency and outcomes of breakthrough infections. Using both pharmacologic records and the COVID-19 registry at the Cleveland Clinic we have systematically examined a large population of IMIDs patients who have received BCDTs and were vaccinated against SARS-CoV-2, and have described the frequency of breakthrough infections and their outcomes.

## Methods

### Patient Identification and Data Extraction

To identify breakthrough patients of interest, all pharmacy records from within the Cleveland Clinic were electronically searched for patients undergoing B cell depletion with approved monoclonal antibodies in the year 2020. Records with ICD codes for IMIDs (see Supplement Table 1) but not malignancies were identified. Using the Cleveland Clinic COVID-19 Research Registry which tracks vaccine and COVID-19 data across our system, we cross-referenced these patients to identify those who were vaccinated at least once and experienced breakthrough infection. Only infections confirmed by PCR or rapid test at any time after the first vaccination were included. In addition, any patients identified through routine care who fulfilled the above criteria were included. This additional strategy allowed identification of breakthrough patients who may have received BCDT outside of our health system or who were diagnosed outside our health care system but who were nevertheless under the care of our providers.

Additional data on demographics (age, gender, race), comorbidities (BMI > 30, heart disease, pulmonary disease, chronic kidney disease, malignancy, smoking history), associated immunosuppressive medications, prednisone use and dosage, timing and duration of BCDT, vaccine type and number of doses, whether or not the patient received outpatient monoclonal antibody therapy for COVID-19, and clinical outcomes were extracted by individual chart review from patient electronic medical records. Each breakthrough infection was classified as complete [i.e. 14 or more days following the second vaccine (or first Johnson & Johnson)] or incomplete.

### Outcome Assessment

Our primary outcome was disease severity; each patient was classified on the 8 point NIH COVID-19 ordinal scale^16^ (Table 1). Patients were classified to their highest level of disease severity and grouped as mild (infections managed outside of the hospital and not requiring supplemental oxygen - Groups 1 and 2) or severe (those requiring hospitalization – Groups 3-8).

**Table 1.**
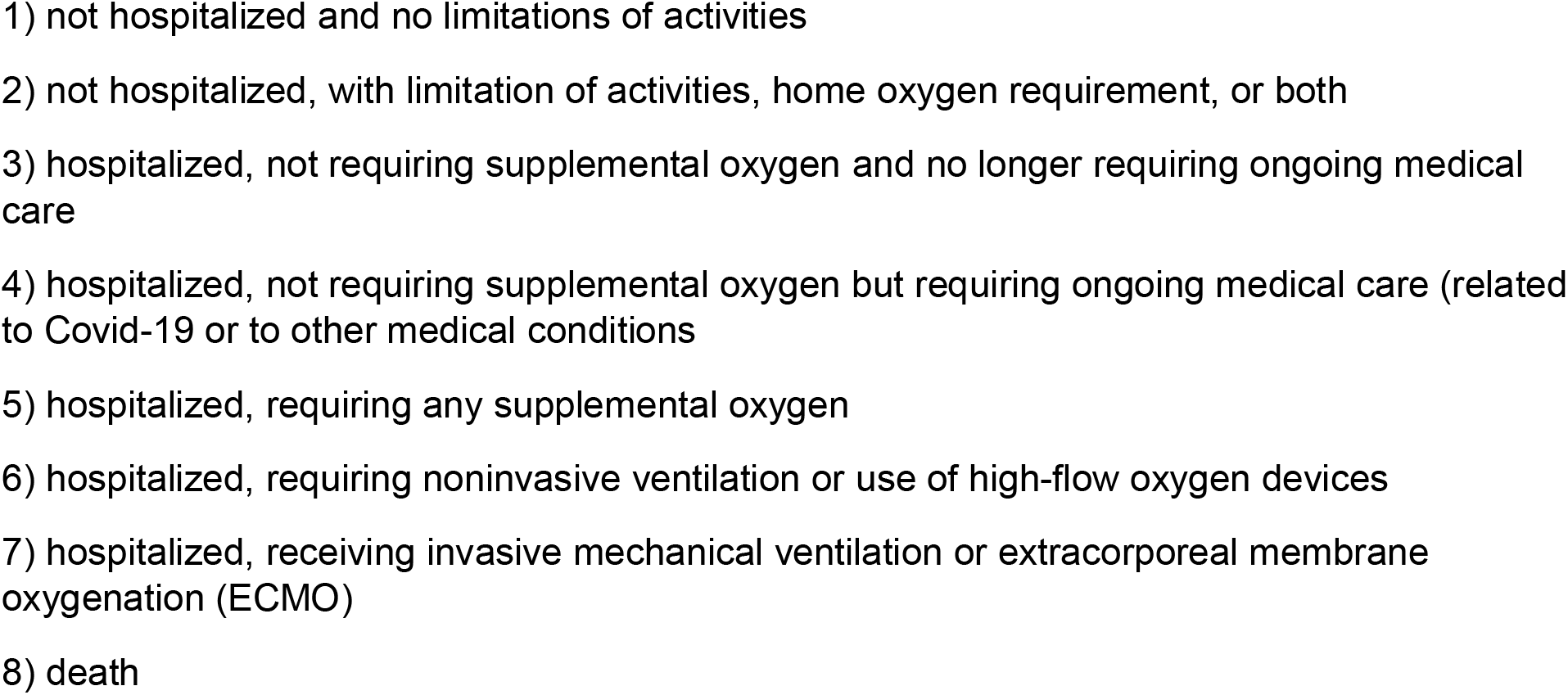
Eight-point ordinal Scale for COVID-19.

### Influence of Delta Variant on Infection Incidence

To account for changes in COVID-19 breakthrough infection rates attributable to the Delta variant, we used June 20, 2021, as the date by which to stratify the follow-up period into pre– or post–Delta phases. The date was based on the CDC report of Delta being the dominant SARS-CoV-2 strain (>50%) in the US^17^. The end date of the study was December 15^th^ 2021, selected based on the date that the Cleveland Clinic microbiology laboratory monitoring the appearance of SARS-CoV-2 variants in Northeastern Ohio identified it as the time point when Omicron became and persisted as the dominant variant.

The Cleveland Clinic Institutional Review Board approved this study.

### Statistical analysis

Continuous variables were summarized using mean and standard deviation, or median and interquartile range (IQR) when appropriate. Categorical variables were summarized using counts and frequencies. For patients who received monoclonal antibody versus those who did not, two-sample t-tests (or Wilcoxon tests) and Pearson Chi-square tests (or Fisher’s exact tests) were performed. Person-time (at risk) accrued from the date of the first dose of the vaccine to the date of breakthrough infection or Dec 15, 2021, whichever occurred first. Unadjusted Poisson regression was used to calculate the overall incidence rate, and the incidence rates of pre-Delta and post-Delta periods. We then investigated the association between each risk factor and COVID-19 severity outcome using univariate logistic regression. Additionally, we built multivariable logistic regression models to examine the effect of potential risk factors on severity outcome after controlling for confounding variables. Data management and analysis were conducted using R Software (Version 4.0; Vienna, Austria). All tests are two-sided, with an alpha level of 0.05.

## Results

### Clinical Features of the Cohort

The results of our search revealed 1696 patients with IMIDs (as identified via specific ICD 10 codes) receiving one or more doses of approved monoclonal antibodies directed against B cells in the year 2020. These patients also received one or more doses of vaccine against SARS-CoV-2. From these cases 74 were found to have experienced a breakthrough infection from the time of their first vaccine through December 15, 2021. The details of these patients are displayed in Table 2. The patients were nearly equally distributed between rheumatic and neuroinflammatory disease with multiple sclerosis the single most common diagnosis. Thirty-four (45.9%) patients were on an additional immunosuppressive drug with prednisone being the most common (35%). Thirteen patients (17.6%) were on one or more additional disease-modifying anti-rheumatic drugs including six on methotrexate, four on azathioprine and four on mycophenolate. The majority of patients (51, 68.9%) had one or more comorbidities.

**Table 2:** Patient Characteristics.

|  |  |
| --- | --- |
| **Median age** | **53 years** |
|  | N (%) |
| **Female** | 46 (62.2) |
| **White race** | 62 (83.8) |
| **Diagnosis** |  |
| Inflammatory CNS | 34 (45.9) |
| Vasculitis | 20 (27) |
| RA | 9 (12.2) |
| Other<sup>1</sup> | 11 (14.9) |
| **Comorbidities** |  |
| BMI 30+ | 35 (47.3) |
| Heart Disease <sup>2</sup> | 36 (48.6) |
| Pulmonary <sup>3</sup> | 13 (17.6) |
| CKD | 8 (10.8) |
| Malignancy | 6 (8.1) |
| **# of Comorbidities** |  |
| 0 | 23 (31.1) |
| 1 | 24 (32.4) |
| 2 | 9 (12.2) |
| 3+ | 18 (24.3) |
| **Vaccine** |  |
| Pfizer | 45 (60.8) |
| Moderna | 23 (31.1) |
| J & J | 6 (8.1) |
| **BCDT** |  |
| < 1 year | 20 (27) |
| 1-3 years | 20 (27) |
| 3+ years | 34 (45.9) |
| **Immunosuppression** |  |
| GC <10 mg/day | 20 (27) |
| GC ≥ 10 mg/day | 6 (8.1) |
| 1 DMARD | 12 (16.2) |
| 2 DMARD | 1 (1.3) |
| **Time between BCDT & vaccine #1** |  |
| <3 months | 22 (32.8) |
| 3-6 months | 36 (53.7) |
| >6 months | 9 (13.4) |
| **Time between BCDT & COVID-19 diagnosis** |  |
| <3 months | 36 (48.6) |
| 3-6 months | 21 (28.4) |
| >6 months | 17 (22.9) |
| **Received mAb therapy for COVID-19 <sup>4</sup>** | 21 (31.3) |

**Abbreviations** RA: rheumatoid arthritis; CNS: central nervous system; CKD: chronic kidney disease; BCDT: B-cell depleting therapy; GC: glucocorticoid; DMARD: disease-modifying antirheumatic drug (includes hydroxychloroquine, methotrexate, azathioprine, mycophenolate); mAb: monoclonal antibody

In terms of vaccination history and status, 45 patients received the Pfizer mRNA vaccine: 2 patients received only one dose before breakthrough, 39 had two doses, and four patients had an additional 3^rd^ dose. Twenty-three patients received Moderna mRNA vaccine with only 1 having a single vaccination before breakthrough, 17 having two doses, and five an additional 3^rd^ dose before infection. Six patients received a single Johnson & Johnson vaccination before breakthrough. Overall, of those experiencing breakthrough infections defined as 14 days past the second vaccination (or single Johnson & Johnson), 6 of 74 were incomplete in their vaccination status while 68 were either completely vaccinated or boosted.

### Outcomes

Clinical outcomes for the entire group revealed that 48 patients had mild outpatient-managed disease with 33 (44.6%) in group 1 and 15 (20.3%) in group 2. The remaining 26 (35.1%) patients were hospitalized with 4 (5.4%) not requiring supplemental O_2_ (groups 3 and 4) and the remaining all requiring some level of O2 support with 11 (14.9%) (Group 3) requiring any O_2_, and 5 (6.8%) (Groups 6 or 7) requiring high flow O_2_ or mechanical ventilation. There were 6 (8.1%) deaths. In terms of risk factors for severe outcomes we examined the groups based on clinical grade by ordinal scale, separating the patients into two groups: those with mild disease (groups 1 and 2), and those classified as severe including all patients requiring hospitalization (groups 3-8). Univariate and multivariate analyses (Table 3) comparing mild (ordinal scale groups 1 and 2) to severe (ordinal scale groups 3-8) revealed that only the presence of two or more comorbidities (present in 21 of the 74 breakthrough patients) was associated with disease severity (P<.004; P = 0.031, respectively). Vaccine associated variables (i.e. complete, incomplete, boosted) had no association with severe outcomes nor did the use of concomitant immunosuppressive therapies. Analysis of BCDT-associated variables including duration of therapy and the time interval from their most recent BCDT treatment to their vaccination demonstrated no statistical effect on risk of severe outcomes in either univariate or multivariate analysis. Finally, four of our breakthrough patients had had a single prior episode of COVID-19 before receiving their first vaccine which had no association with severe outcomes in univariate analysis (data not shown).

**Table 3:**
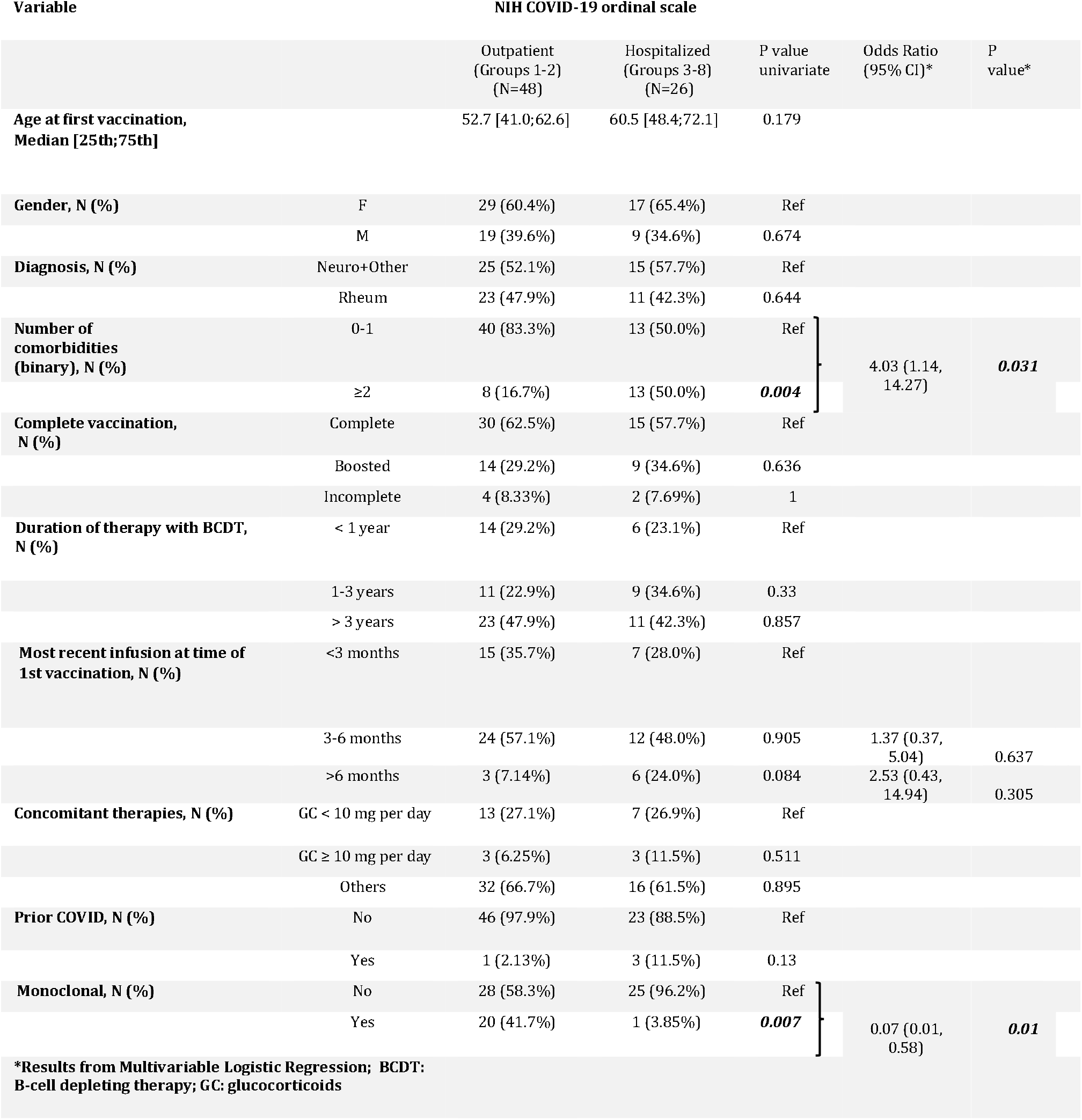
Univariate and multivariate analysis of clinical variables with potential impact on COVID-19 severity.

### Incidence

Of 1696 patients with any form of vaccination, 74 had breakthrough infection for a raw incidence of 4.36%; 68 (91%) occurred after complete vaccination (i.e. 14 days after the final administration). We also examined the crude incidence of COVID-19 breakthrough as a function of the pre-Delta and post-Delta periods in 2021. Prior to June 20 (the date CDC identifies as the onset of the Delta surge) there were 12/74 cases identified while after June 20 there were 62, supporting a seeming acceleration of breakthrough infections with the Delta variant. The total person-time of vaccine exposure for this cohort (1696 patients with at least one vaccine) was 14302.67 months. As a result, the time-adjusted incidence rate of breakthrough infection in the entire group is 5.19 per 1000 person-months (95%CI, 4.13-6.52). The time-adjusted incidence rate of breakthrough infection in the pre-Delta period is 2.48 per 1000 person-months (95% CI, 1.41-4.36). The incidence rate in the post-Delta period is 6.59 per 1000 person-months (95% CI, 5.14-8.45).

### Effects of outpatient therapy with monoclonal antibodies for COVID-19

The use of anti-SARS-CoV-2 monoclonal antibody therapy (casarivimab and indevimab) within 10 days of symptom onset as outpatient therapy for COVID-19 was employed in 21 patients. No patients in our cohort received any other type of monoclonal antibody treatment for COVID-19. In the multivariable model, after controlling for both number of comorbidities and most recent time of first vaccination administration, this therapy was associated with more favorable outcomes with only one patient requiring hospitalization (without O_2_ support) and no deaths (ordinal scale group 3) (p=0.007) (Table 3). The results of this intervention and effects on highest level of ordinal scale severity is displayed graphically in Figures 1a and b. To explore the possibility that those treated with monoclonal antibodies differed in some manner that could potentially contribute to their markedly different clinical outcomes we compared a select number of clinical characteristics (age, number of comorbidities, concomitant immunosuppressive therapies, duration of BCDT) between those who received monoclonal antibody therapy and the entire breakthrough cohort; none were statistically significant (Table 4).

**Figure 1.**
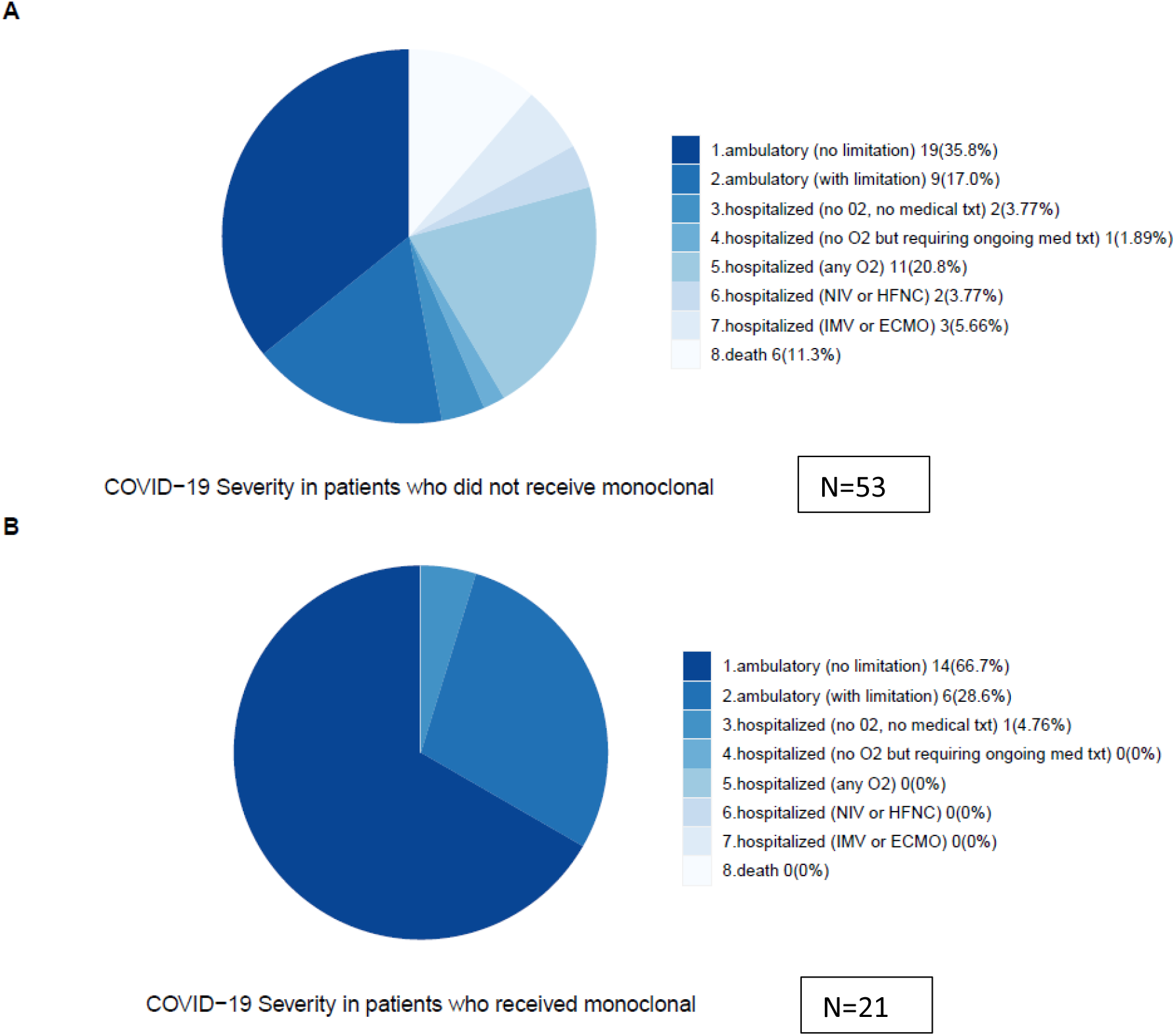
Pie chart for COVID-19 severity outcome distribution based on whether or not received outpatient monoclonal antibody therapy.

**TABLE 4:** Comparisons of clinical features between patients receiving monoclonal antibody therapy versus patients not receiving monoclonal antibody therapy.

|  |  | Total(N=74) | Not on monoclonal(N=53) | On monoclonal(N=21) | Pvalue |
| --- | --- | --- | --- | --- | --- |
| Age at first vaccination, Median [25th;75th] |  | 55.2 [41.7;65.4] | 54.9 [44.0;65.5] | 55.5 [37.6;65.1] | 0.679 |
| Gender, N (%) | F | 46 (62.2%) | 34 (64.2%) | 12 (57.1%) | 0.768 |
|  | M | 28 (37.8%) | 19 (35.8%) | 9 (42.9%) |  |
| Concomitant Therapies 2, N (%) | Glucocorticoids < 10 mg per day | 20 (27.0%) | 11 (20.8%) | 9 (42.9%) | 0.129 |
|  | Glucocorticoids > 10 mg per day | 6 (8.11%) | 4 (7.55%) | 2 (9.52%) |  |
|  | Others | 48 (64.9%) | 38 (71.7%) | 10 (47.6%) |  |
| Number of Comorbidities (binary), N (%) | 0-1 | 53 (71.6%) | 35 (66.0%) | 18 (85.7%) | 0.160 |
|  | >=2 | 21 (28.4%) | 18 (34.0%) | 3 (14.3%) |  |
| Duration of therapy with B cell agent, N (%) | < 1 year | 20 (27.0%) | 15 (28.3%) | 5 (23.8%) | 0.783 |
|  | 1-3 years | 20 (27.0%) | 15 (28.3%) | 5 (23.8%) |  |
|  | > 3 years | 34 (45.9%) | 23 (43.4%) | 11 (52.4%) |  |

## Discussion

The current study examines a large cohort of patients exposed to BCDT in 2020, a time frame that began nearly 12 months before introduction of the SARS-CoV-2 vaccine, all of whom received one or more vaccinations for COVID-19 and developed breakthrough infection. Breakthrough occurred in 74 (4.4 %) or nearly 1 in 20. Our data clearly show that breakthrough infection in this population is associated with severe outcomes, as 35.1% required hospitalization, 15% required critical care, and 8% died. Thus we confirm that IMIDs patients on BCDTs appear vulnerable to breakthrough infections and, most importantly, are attended by severe outcomes.

While patients with IMIDs on BCDTs have been recognized to be vulnerable to severe COVID-19 infections,^3,4,6,7^ relatively scant data exist regarding their risks for and outcomes of breakthrough infections^2,18^. The current cohort of breakthrough patients consisted of nearly equal numbers of patients with rheumatic and neuro-inflammatory disease and most (91.9%) were either fully vaccinated or boosted, with only 6 (8.1%) with incomplete vaccination. Of note is that the group as a whole was heavily exposed to BCDTs with 27.0% on therapy for one to three years and 45.9% for more than three years. More importantly, 76.5% patients received their last monoclonal antibody therapy less than six months before their first vaccine, a time point that several studies have demonstrated is critical for any reasonable chance of developing a humoral response^10,12,14^.

Although our study lacked comparator groups of healthy or immunocompromised patients not on BCDTs to appraise breakthrough frequency and severity, indirect comparisons to previously reported studies provide some perspective for interpreting our findings. Sun et. al.^1^, in a large study of breakthrough infections in immunocompromised patients, retrospectively examined 664,772 patients in the National COVID Cohort Collaborative with a variety of disorders sharing varying degrees of immune dysfunction. This study was not broken down by therapies. The authors reported an overall rate of serious disease (as determined by need for hospitalization) of 20.7%, far less than the 35% observed in our cohort. Comparing COVID-19 severity in our group to CDC estimates of hospitalization for breakthrough infections in the general population for the week ending December 18, 2021 revealed hospitalization rates for 18-49 year olds was 1.9%, 50-64 years old 4%, and 65+ years and older 12.4%, all far below our observed rates of hospitalization ^19^.

In terms of our overall rate of breakthrough infection, a study by the CDC in the general population estimated a breakthrough rate of 2.8% per six month period on September 30, 2021 and does not appear markedly different from our observed crude rate of 4.4% over nearly 12 months of observation^20^. In terms of our time adjusted incidence rate of 5.19 per 1000 person-months it appears comparable to the overall rate of 5.0% (95%, CI 2.9-2.9) in immunocompromised patients in the study by Sun et. al.^1^

Our study also examined the impact of the Delta variant on breakthrough incidence and found that the majority of cases were identified during the period of the Delta surge, with 62 of the 74 cases diagnosed between June 20 (the date the CDC marked as the onset of the Delta surge^1^) and December 15 (the end of our study and the end of the Delta surge in Northeastern Ohio). This observation is consistent with the study of Sun et. al., ^1^ who described a tripling of incidence in the period following June 20 until the end of their study period of September 16^th^, 2021. In terms of time adjusted incidence in the pre-delta period our study of 2.48 per 1000 person-months (95% CI, 1.41-4.36) is comparable to the 2.9% per 1000 person months (95% CI, 2-9-29) in the study of Sun et. al.^1^ but our incidence rate in the post-Delta period is 6.59 per 1000 person-months (95% CI, 5.14-8.45) is less than the 9.6 per 1000 person-months in the overall group of immunocompromised patients in this same study. The reasons for this are unclear but the study populations while sharing the immunocompromised state were not the same and our search methodologies were different.

Our findings of severe outcomes of COVID-19 in breakthrough infections in patients being treated with BCDTs is important for understanding the implications of the evolving picture of vaccine responsiveness in this population. From early on it has been appreciated that patients on agents such as rituximab and ocrelizumab display severe deficits in their capacity to mount humoral responses to a variety of vaccines and more recently to SARS-CoV-2^10^. In the last two years, however, several more detailed studies have revealed a surprisingly robust T cell response following m-RNA vaccines ^14,15,21^, suggesting the possibility that the sum total of clinical immunity (i.e. protection from the sequelae of SARS-C0V-2 infection) may be salvaged by augmented cell mediated responses. Unfortunately our data clearly indicate that in general this does not appear to be the case, as these patients remain vulnerable to severe disease and poor outcomes.

Our observation that patients receiving monoclonal antibodies did extremely well, supported by the fact that only a single patient was hospitalized without requiring oxygen therapy, is important. While monoclonal antibodies have been used extensively their utility at reducing the need for acute care and death is based on clinical trials in patients largely at increased risk based on age and concomitant diseases as opposed to a small minority of patients with immunocompromising conditions and none that we are aware of explicitly recruiting patients on BCDTs such as found in our study^22^. Our comparative analysis of those treated with outpatient monoclonals versus the larger group of breakthrough patients for select clinical characteristics which may have biased the response revealed no differences, supporting the efficacy of this therapy. The seeming underutilization of COVID-19 monoclonal antibody treatment in our patients is disappointing, with only 21 of 74 being treated. Possible explanations include periods of time when these therapies have had limited availability or when patients failed to connect with their practitioners within the ten day window of eligibility. Also plausible and anecdotally noted in our chart review were uncertainties on behalf of the patients who were diagnosed promptly as to which provider to contact (i.e. primary care or specialist), at times leading to delays caused by caregivers who were unfamiliar with rapidly changing care pathways. Moving ahead it is important to note that monoclonal antibody therapies demonstrate highly strain-specific antiviral activity^23^ and accordingly the results from the use of a single product (casarivimab/indevimab) in our study, while highly effective for Delta and pre-Delta variants, cannot necessarily be extrapolated to either Omicron, which displays differential sensitivity, or future variants.

Our study has several limitations. First, we have no direct comparator group of immunocompromised patients based on therapy, as it would be of interest to compare severity and outcomes to other patients on different immunosuppressive therapies. For now unfortunately large studies such as these have not been reported. Secondly, it is likely that unknown cases of mild or even asymptomatic infection may have been unreported or missed and certain data fields extracted from the chart review may have been missing especially from patients receiving their BCDT outside of our health care system. Thirdly, while we chose to examine patients given BCDTs in the year 2020, we did not account for ongoing BCDT through the end of the study which may have further contributed to immunosuppression. A recent study of rituximab in vasculitis patients demonstrated that antibody levels to S protein fell by greater than 50% within 4 weeks of drug administration ^24^. Finally, it would clearly be of interest to examine breakthrough infections in concert with assessing the status of patients’ integrated immune responses by assessing serologic responses, especially anti-Spike antibody titers, which have been associated with breakthrough infections in IMIDs patients ^25^, as well as B cell numbers and cell mediated immune responses. Unfortunately this was not possible in this retrospective study where such data were not gathered or were missing on the vast majority of patients.

In terms of practical implications our study should serve to highlight the plight of this important segment of the immunocompromised patient population who are likely to face ongoing and formidable risks despite aggressive vaccination if, as many observers predict, an ensuing endemic phase of the pandemic lies ahead with future variants of unknown pathogenicity. For now enhanced non-pharmacologic measures (masking, social distancing, etc.) will remain important; expanded access to pre-exposure prophylaxis with monoclonal antibodies effective against prevalent variants and access to emerging antiviral therapies will be vital^26,27^. Enhanced education of both patients and the practitioners who care for them to increase their awareness and utilization of current and future outpatient therapies is urgent.

## Data Availability

All data produced in the present work are contained in the manuscript

